# Optimisation of SARS-CoV-2 culture from clinical samples for clinical trial applications

**DOI:** 10.1101/2024.03.25.24304829

**Authors:** Dominic Wooding, Kate Buist, Alessandra Romero-Ramirez, Helen Savage, Rachel Watkins, Daisy Bengey, Caitlin Greenland-Bews, Caitlin R Thompson, Nadia Kontogianni, Richard Body, Gail Hayward, Rachel L Byrne, Susan Gould, CONDOR Steering Group, Christopher Myerscough, Barry Atkinson, Victoria Shaw, Bill Greenhalf, Emily Adams, Ana Cubas-Atienzar, Saye Khoo, Tom Fletcher, Thomas Edwards

## Abstract

Clinical trials of SARS-CoV-2 therapeutics often include virological secondary endpoints to compare viral clearance and viral load reduction between treatment and placebo arms. This is typically achieved using RT-qPCR, which cannot differentiate replicant competent virus from non-viable virus or free RNA, limiting its utility as an endpoint. Culture based methods for SARS-CoV-2 exist; however, these are often insensitive and poorly standardised for use as clinical trial endpoints.

We report optimisation of a culture-based approach evaluating three cell lines, three detection methods, and key culture parameters. We show that Vero-ACE2-TMPRSS2 (VAT) cells in combination with RT-qPCR of culture supernatants from the first passage provides the greatest overall detection of Delta viral replication (22/32, 68.8%), being able to identify viable virus in 83.3% (20/24) of clinical samples with initial Ct values <30. Likewise, we demonstrate that RT-qPCR using culture supernatants from the first passage of Vero hSLAM cells provides the highest overall detection of Omicron viral replication (9/31, 29%), detecting live virus in 39.1% (9/23) of clinical samples with initial Ct values < 25. This assessment demonstrates that combining RT-qPCR with virological end point analysis has utility in clinical trials of therapeutics for SARS-CoV-2; however, techniques may require optimising based on dominant circulating strain.

## INTRODUCTION

The design of COVID-19 therapeutic clinical trials and appropriate selection of viral endpoints is crucial to determining treatment efficacy^1^. A variety of endpoints have been identified by systematic reviews of SARS-CoV-2 trial endpoints, including death, recovery, need for intensive care, hospital discharge, oxygenation, critical illness assessment tools, and viral load assessment^1–3^ and often multiple secondary endpoints are selected for analysis.

The use of viral load assays as an endpoint has also underpinned the early-phase evaluation of antiviral activity, such as remdesivir^4^, molnupiravir^5^ and Nirmatrelvir^6^. The gold standard method for detecting and quantifying SARS-CoV-2 in clinical samples is quantitative reverse-transcriptase PCR (RT-qPCR) detection of viral RNA^7^. However, RT-qPCR cannot distinguish between infectious virus and non-infectious degraded RNA fragments that persist after neutralisation by the immune system and, therefore, may over-estimate the presence of infectious virus and under-estimate efficacy of the assessed pharmaceutical. Previous studies have shown a correlation between viral load by RT-qPCR during SARS-CoV-2 infection and culture positivity, with culture positivity being used as an estimate of infectiousness^8^.

Since these RNA-based detection assays do not discriminate between replication-competent virus and remnants of genetic material, an alternative approach is to use viral culture as a proxy for antiviral efficacy. Whilst viral culture is less sensitive than RT-qPCR, it has the advantage of confirming viral infectivity and therefore transmission potential^9,10^. Culture based methods for detecting SARS-CoV-2 are not well standardised and numerous cell lines and culture conditions have been reported^11^. SARS-CoV-2 cell line susceptibility is influenced by many factors including cell tropism, receptor expression levels, virus replication kinetics, and the epidemiological and clinical features of the virus^12^. Globally, the Vero E6 African green monkey kidney cell line is commonly used as a readily available cell line that is permissive for infection by many viruses. Vero E6 cells do not express all SARS-CoV-2 key surface molecules, and viral entry and fusion mainly occurs via non-specific endocytic mechanisms^13^. Alternative Vero cells have been modified to more closely resemble the human epithelia., e.g. Vero hSLAM cells that express the human signalling lymphocytic activation molecule (SLAM)^14^, and VAT cells -Vero E6 expressing both human angiotensin-converting enzyme 2 (ACE2), the major receptor of SARS-CoV-2, and transmembrane serine protease 2 (TMPRSS2), which cleaves the viral S protein priming it for cellular infection^15^. Both viral growth kinetics and changes to cell morphology (i.e., cytopathic effects; CPE) can vary between these different cell lines thereby impacting the outcome of tests used to assess culture positivity^16^. For example, a 2020 study found that cell culture supernatants from Vero E6 cells expressing TMPRSS2 had more than 100 times more viral RNA copies than Vero E6 cells not expressing this protein^17^.

There are various methods for assessing culture positivity, including the use of microscopy to detect CPE caused by viral infection^18^ ^19^, plaque assays to quantify infectious virus in the culture supernatants^20^, and RT-qPCR to detect increases in viral RNA during culture^21^.

To develop a virological endpoint for trials of SARS-CoV-2 therapeutics, it is imperative that all these methodological variables are assessed with the process optimised for maximal sensitivity to enable accurate determination of individuals with infection-competent SARS-CoV-2 in the nasopharynx. In this paper we describe the optimisation of a viral culture assay for detecting infectious virus with assessment of three potential detection methods and three different cell lines.

## METHODOLOGY

### Variables and experimental design

Five variables were identified for optimisation (Table 1), and within each variable a set of parameters were assessed (Fig.1). Variables were tested using samples from the United Kingdom (U.K.) Delta outbreak and, once the methodology was optimised, the procedure was assessed with samples from the Omicron BA.1 outbreak.

**Figure 1.**
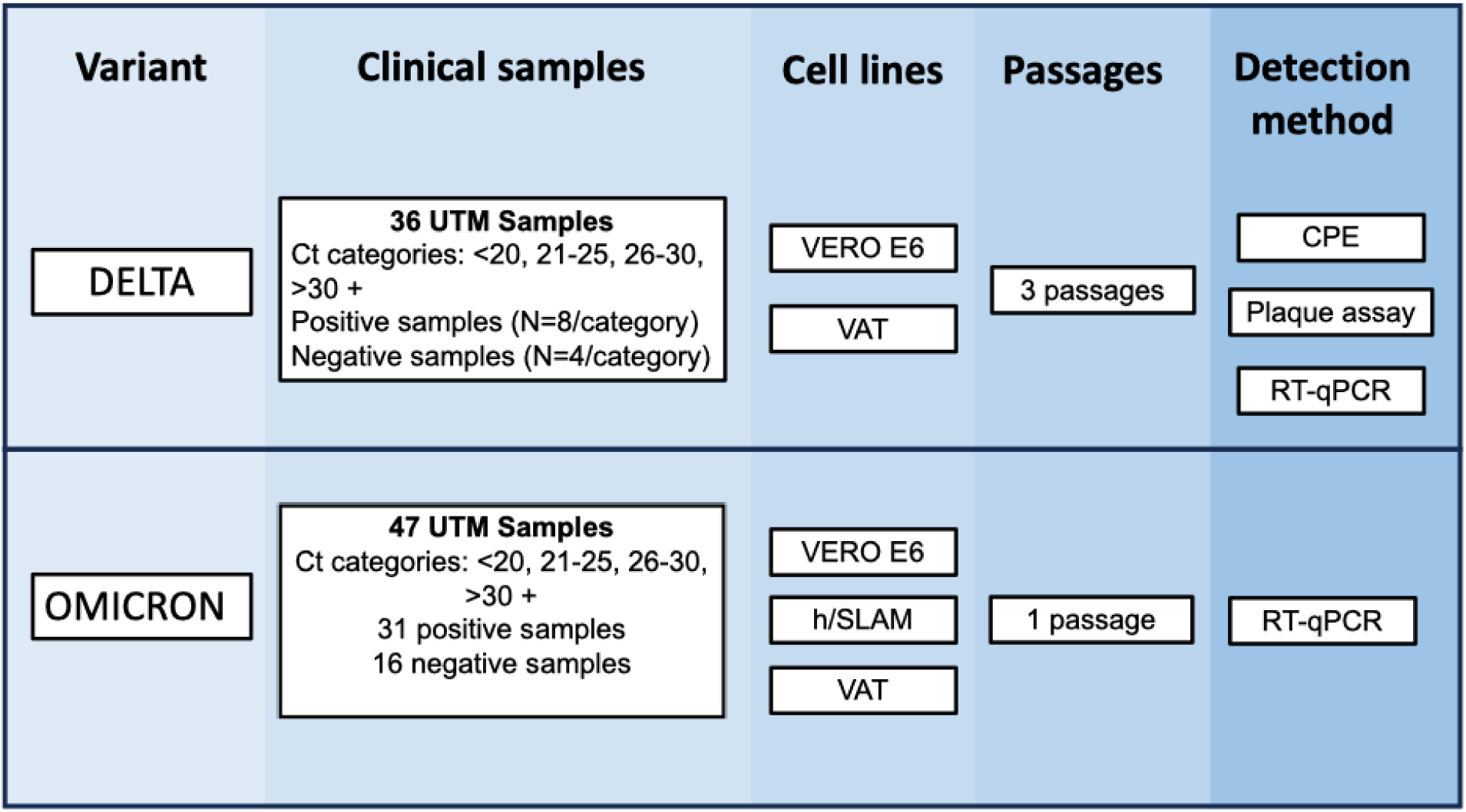
Experimental design for the optimisation of SARS-CoV-2 culture from clinical samples. Scheme of the initial experiments performed using different parameters before the optimization of detection methods using clinical samples. Samples during the Delta and Omicron outbreak were cultured in Vero E6, VAT and/or Vero hSLAM for 3 days. Supernatants were collected to perform RT-PCR for Delta and Omicron respectively and CPE imaging and plaque assays were performed for Delta only.

**Table 1:**
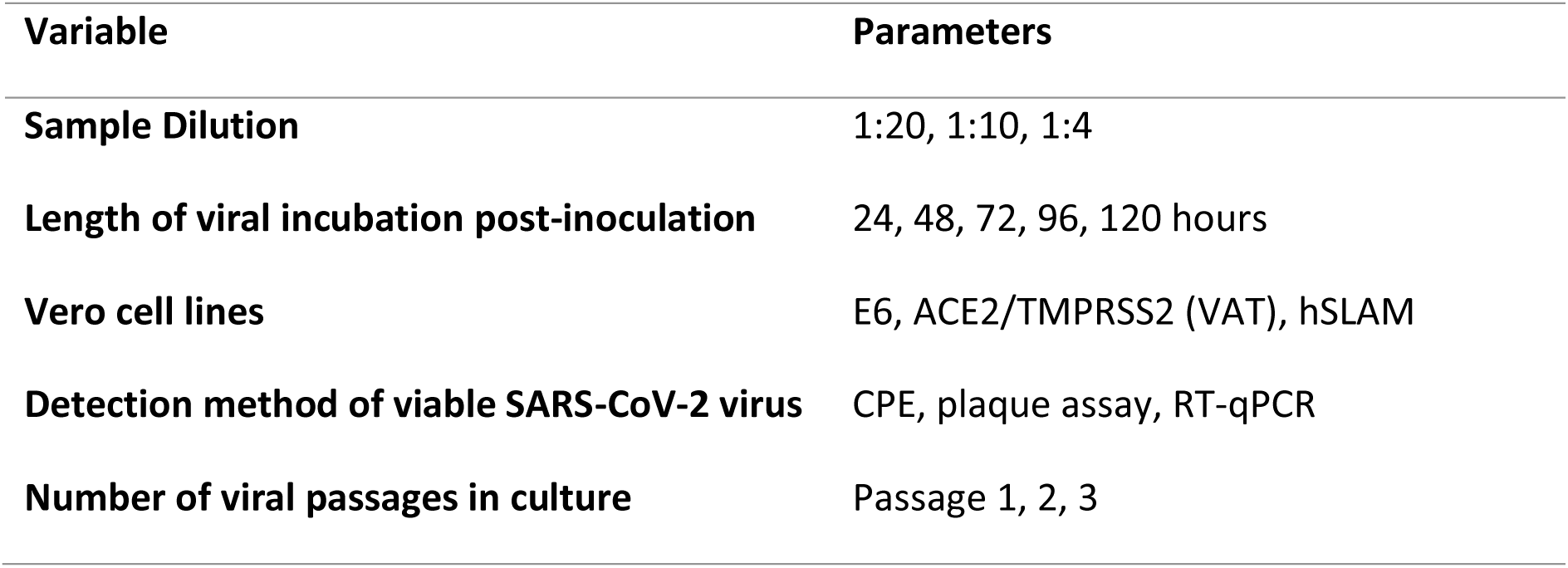
Parameters chosen for the optimisation of the viral culture assay. CPE = cytopathic effect; RT-qPCR = quantitative reverse transcriptase PCR.

| Variable | Parameters |
| --- | --- |
| Sample Dilution | 1:20, 1:10, 1:4 |
| Length of viral incubation post-inoculation | 24, 48, 72, 96, 120 hours |
| Vero cell lines | E6, ACE2/TMPRSS2 (VAT), hSLAM |
| Detection method of viable SARS-CoV-2 virus | CPE, plaque assay, RT-qPCR |
| Number of viral passages in culture | Passage 1, 2, 3 |

#### 1. Cell culture

Vero C1008 [Vero 76, clone E6, Vero E6] (ECACC 85020206) (Vero E6 cells) and Vero hSLAM (ECACC 04091501) (hSLAM cells) were obtained from the European collection of authenticated cell cultures (ECACC). Vero E6-ACE2-TMPRSS2 cells (VAT cells) were donated from Professor Wendy Barclay (Imperial College London, UK), and their development is detailed in a publication from 2021^15^. For the duration of the experiment, Vero E6 cells were maintained in T125 cell culture flasks containing 25ml of Dulbecco’s Modified Eagle Medium (DMEM, Gibco, USA) plus 10% foetal bovine serum (FBS, Gibco, USA) and 1% Penicillin/ Streptomycin solution (Gibco, USA) (D10 media) at 37°C and 5% CO_2_. VAT cells were maintained as described above, with a modified growth medium: DMEM plus 10% FBS, 2% 50mg/ml Geneticin (Gibco, USA), 1% 100X Non-essential Amino Acids (ThermoFisher, USA), and 0.4% 50mg/ml Hygromycin B (Invitrogen, USA)). hSLAM cells were maintained as above but with the following growth medium: Minimum Essential Media GlutaMAX™ (MEM, Gibco, USA) plus 10% FBS and 2% 50mg/ml Geneticin. Adjustments in the FBS content of these maintenance media to 4% (D4 media) or 2% (D2 media) were required at various points in the culture process, detailed below. Once cells reached 100% confluency (every 3-4 days), the media was removed, cells were washed with 10ml phosphate buffered saline (ThermoFisher, USA), and 2ml of Trypsin-EDTA solution (Gibco, USA) was added to dissociate cells from the flask. To inactivate the trypsin, 8ml of D10 media was added, and cells were pipette-mixed to separate clumps. Culture flasks were re-seeded at a 1:10 ratio which completed one passage. Cells were maintained for no more than 30 passages, so that the integrity of the cell lines was not compromised.

The optimal sample volume and number of days growth of virus per passage was determined with the Delta variant of concern (VOC) as stated in the Supplementary material section (Supplementary material a. Sample dilution and b. SARS-CoV-2 growth curves).

#### 2. Plaque assay

For establishing plaque assay plates, dissociated cell suspension was counted using a Primovert inverted light microscope (ZEISS, Germany) and a disposable C-chip Haemocytometer (NanoEnTek, South Korea), before diluting with D10 media to a concentration of approximately 250,000 cells/ml. Cells were seeded on 24-well culture plates (ThermoFisher, USA) with approximately 500μl of 250,000 cells/ml per well. Plates were incubated at 37°C + 5% CO_2_ for 18 hours to produce a confluent monolayer of cells.

Media was discarded from all wells, and 190µL of fresh D2 media was added. 10µL of sample was added to the appropriate wells and incubated at 37°C + 5% CO_2_ for 1 hour. An overlay solution (prepared by combining equal parts of 2.2% (w/v) Agarose solution (Sigma-Aldrich, USA) and D4 media) was added at 500 µL/well and the plate was incubated at 37°C + 5% CO_2_ for a further 72 hours. Approximately 1ml/well of Formaldehyde solution (37% w/v) (Merck, Germany) was added, and plates were incubated at room temperature for a minimum of 1 hour. The contents of each well were discarded into vermiculite, and plates were stained with crystal violet solution (0.25% w/v in distilled water) for 1 minute, rinsed twice with tap water before air drying and plaque enumeration.

#### 2. RT-qPCR

For the RT-qPCR assays from Delta and Omicron culture supernatants, RNA was extracted from supernatants using the QiAamp96 Virus Qiacube HT kit (Qiagen, Germany) and RT-qPCRs were run following manufacturer’s instruction using TaqPath COVID-19 RT-PCR on a QuantStudio 5 (ThermoFisher, USA). Fluorescence was recorded in the FAM, VIC, ABY and JUN channels for the SARS-CoV-2 ORF1ab, N, and S gene targets, plus MS2 RT-qPCR internal control target respectively. The N gene value was then selected as the most stable target to stratify the samples (lower mean Ct and standard deviation at 10 genome equivalent copies (GCE)/reaction^22^). Four and 16 RT-qPCR-confirmed negative samples were also selected as controls in the case of Delta and Omicron clinical samples, respectively. The reproducibility of the RT-qPCR was determined to further inform the Ct difference selected to be indicative of a positive culture by testing 10 replicates from a unique UTM sample at 1X limit of detection (LOD).

### SARS-CoV-2 clinical sample cohort

Aliquots of universal transport media (UTM, UTM-RT, Copan, USA) (nasopharyngeal samples) from a cohort of adult participants with symptoms suggestive of COVID-19 were collected by the ‘Facilitating Accelerated Clinical Evaluation of Novel Diagnostic Tests for COVID-19 (FALCON C-19), workstream C (undifferentiated community testing)’. Ethical approval was obtained from the National Research Ethics Service (reference 20/WA/0169) and the Health Research Authority (IRAS ID:28422, clinical trial ID: NCT04408170). Samples were stored at – 80°C and thawed for the first time for this study.

#### 1. Initial evaluation

The major optimisation experiment was carried out using the Delta variant to compare three chosen detection methods: plaque assay, observable CPE, and comparison of RT-qPCR cycle threshold ^23^ values before and after viral culture, in the three cell lines for three passages. All conditions were tested in triplicate. Methods were evaluated for their sensitivity in detecting culture positivity from RT-qPCR positive samples, with the best performer selected for further testing with Omicron samples.

##### a. Delta clinical samples

Samples were selected from those collected between 19^th^ July 2021 and 26^th^ October 2021, based on a >99% frequency of the Delta variant in the UK between these dates^24^. Two different sets of Delta virus positive clinical samples were used for Vero E6 and VAT cell lines, respectively (n=36, each set) due to the availability of the cell lines at different time points and the requirement of avoiding using freeze-thawed samples. However, samples were selected for the study based on the Ct value obtained when tested at the time of sample collection using the TaqPath COVID-19 RT-PCR kit (ThermoFisher, United States). Eight samples were selected for each of the following ranges of Ct values: <20, 21-25, 26-30 and >30. Where possible, to ensure a breadth of Ct values within each range, each category was split in half (e.g., <20 was split into 10-15 and 16-20), with four samples taken for each.

##### b. Impact of viral passage on culture positivity

Cells from each cell line were seeded into 24-well culture plates. D2 media was added at 190µL/well, and 10µL of each UTM sample was added in triplicate. Plates were incubated at 37°C + 5% CO_2_ for 2 hours, before a further 200µL/well of D10 media was added. Plates were incubated at 37°C + 5% CO_2_ for 72 hours to complete the passage. The process was repeated twice, with 10µL/well being transferred to fresh confluent 24-well plates. Each 72-hour incubation was referred to as a viral passage (giving 3 passages total). After each passage, the culture positivity was determined by the methods detailed above.

##### c. Identification of culture positivity

The three detection methodologies used were microscopy for visual inspection of CPE (supplementary methods C), plaque assay and RT-qPCR.

Plaque assays were performed on the 10µL of supernatant taken from each well at each passage described in the “Plaque Assay” section. The presence of plaques indicated that infectious virus was present in the well, which was scored as a positive, otherwise wells were scored as negative. Any plates with detached monolayers after staining were repeated.

The final detection method to assess viral positivity was a comparison of RT-qPCR cycling threshold ^23^ values from a baseline (the original sample diluted 1:40 using D2 media to replicate the dilution factor inherent for viral culture assessment) and the culture supernatant, using the TaqPath RT-qPCR assay. A decrease in Ct value (meaning an increase of viral genetic material) indicates that the virus has successfully replicated and is therefore present and viable. Based on the reproducibility data generated, and that presented in the TaqPath users handbook, we selected a decrease in Ct value of >1 between the baselines and culture supernatant as an indicator of culture positivity.

#### 2. Omicron Variant

##### a. Omicron clinical samples

Clinical samples were collected from 14^th^ December 2021-28^th^ March 2022 based on >99% frequency of Omicron. Samples were also selected based on previous diagnostic results using the Taqpath RT-qPCR kit indicating an S gene target failure^25,26^. In addition to the clinical samples, an Omicron virus stock (1.4×10^5^pfu/ml) was also incubated in triplicate as a positive control. An aliquot from the same virus stock was heat inactivated at 80°C for 1 hour^27^ to generate a negative control and UTM media was used as no virus control.

##### b. Evaluation of optimised culture methodology

The optimized methodology was determined to be a three-day incubation of a single passage, a 1:20 sample dilution, and RT-qPCR based detection. This optimized methodology was validated using 31 positive and 13 negative Omicron samples (Fig. 1). The cell lines Vero E6, VAT and, in addition, hSLAM cells due to high susceptibility for Omicron infection ^28,29^ were used to determine the optimal cell line for this variant. A positive sample (Omicron virus stock), negative sample (inactivated virus stock) and blank (UTM media) were used as controls.

### Statistical Analysis

Data were collated and analysed using R v4.1.1 (R Foundation for Statistical Computing, Vienna). Graphical analysis was undertaken using the ggplot package. The coefficient of variation between replicates and the differences between means on Ct values were performed using an independent two-sample T test using R (version 4.2.1)^30^.

## RESULTS

Thirty-six nasopharyngeal samples collected during the UK Delta outbreak were used to optimise each methodology. Due to the timings of the experiments and a need to avoid freeze-thaw, different samples were used to assess the Vero E6 and VAT cell lines; however, samples were selected with the same Ct ranges (Fig.S2). There was no significant difference between the mean Ct of the two sample sets (Sample set 1 [Vero E6] mean = 25.34; sample set 2 [VAT] mean = 25.48; p-value = 0.934, CI –3.57 – 3.289). However, the mean Ct of the >30 sample category was higher for sample set 1 than for set 2 (35.51 v. 32.46) and significantly different (p-value = 0.025, CI 0.436-5.667). After evaluating the reproducibility of the RT-qPCR assay, the coefficient of variation calculated at 1X LOD was 0.8%.

### Dilution factor

The optimal dilution factor of UTM was assessed to minimise the potential for inhibition or contamination of the cultures without compromising viable virus detection. There was no significant difference in the number of plaques produced with a sample volume of 10µL, 20µL or 50µL within each passage of each cell line (Table S1). However, we did observe signs of contamination in some wells of the 20µL and 50µL volumes, using both microscopy and visual observation of a change in colour of the media. For these reasons, an optimal input sample volume of 10µL (overall dilution = 1:40) was chosen, to reduce the risk of contamination without impacting sensitivity.

### Optimisation results

All SARS-CoV-2 positive clinical samples that were tested in triplicate by RT-qPCR produced a valid result. Despite dilution of input clinical samples, contamination was still observed in 14 (2.4%) of 576 samples replicates (Table S3A), with 2 and 12 wells contaminated in the E6 and VAT cell line, respectively. In addition, 60 of 576 (13.2%) of the original plaque assays failed due to a lack of viable monolayer and were repeated from a frozen aliquot of the same sample, then added to the final data table (Table S3A). All 60 plaque assay replicate failures were in the E6 cell line.

From virus passage 1, the VAT cells resulted in the highest proportion of positive cultures using each detection method (Fig.3). The combination that gave the highest number of positive cultures was RT-qPCR with the VAT cells (68.8%). Whilst further passages improved the positivity rate using Vero E6 cells (34.4%, 43.8% and 43.8% for passages 1, 2 and 3 respectively), the sample positivity in VAT cells decreased across the passages (68.8%, 37.5% and 40.6% for passages 1,2 and 3 respectively).

**Figure 3.**
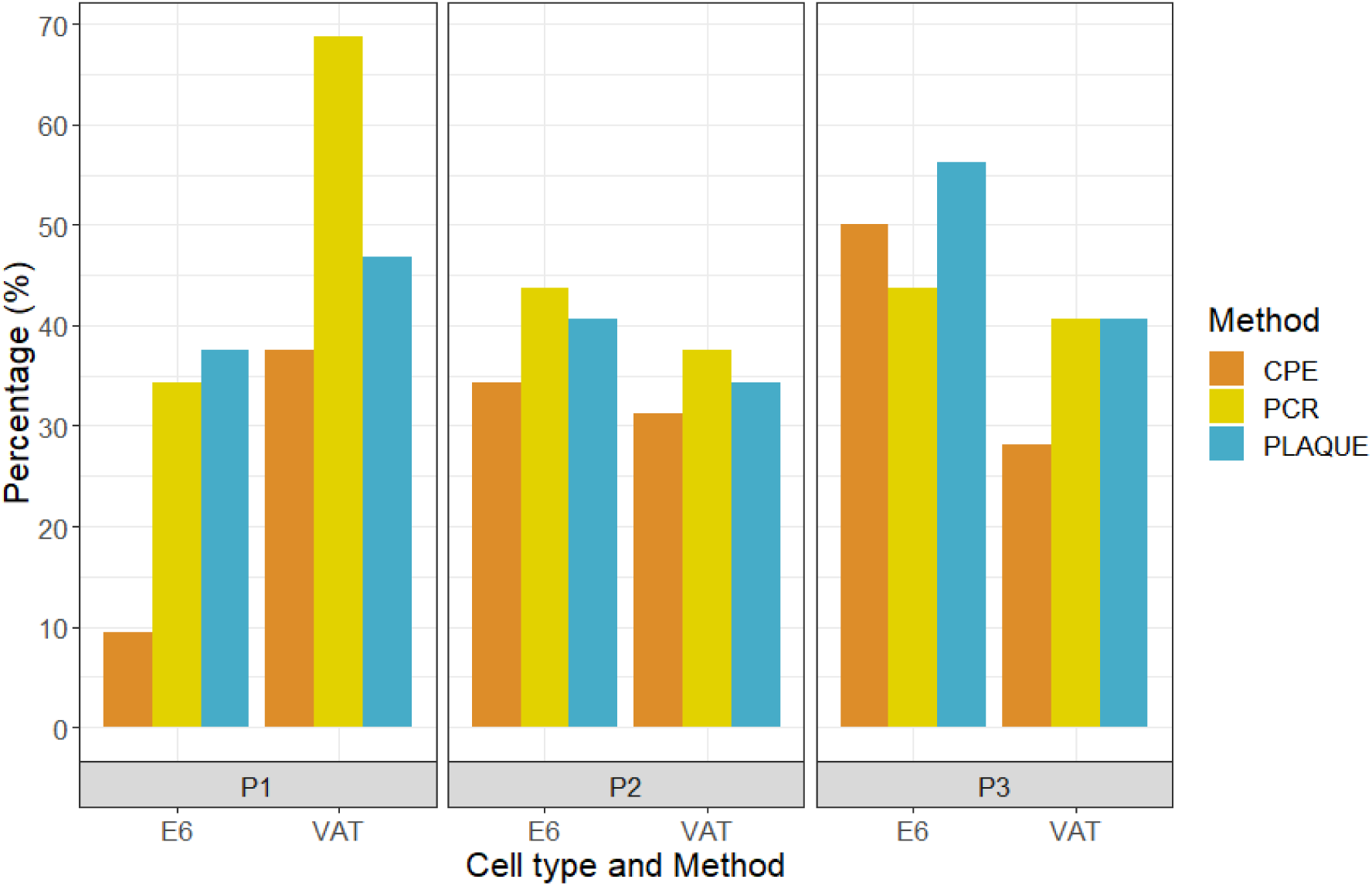
Percentage of positive results by cell type and passage (P1, P2, and P3 refers to passage 1, 2 and 3, respectively) by each detection methodology tested using RT-qPCR positive samples. PCR: RT-qPCR, CPE: cytopathic effect, Plaque: plaque assay.

For each cell line at virus passage 1, microscopy was the least sensitive detection method. For samples in the <20 Ct range, the use of microscopy to detect CPE yielded a positivity rate of 37.5% and 100% in the Vero E6 and VAT cells, respectively. For samples with Ct values in the 20 to40 range, the only positives were found in the VAT cells, with a cumulative positivity of 50% for all samples <Ct 30 (Fig.4A). There were no positive samples found at any range for Vero E6 cells except for Ct<20 (Table S3A).

**Figure 4.**
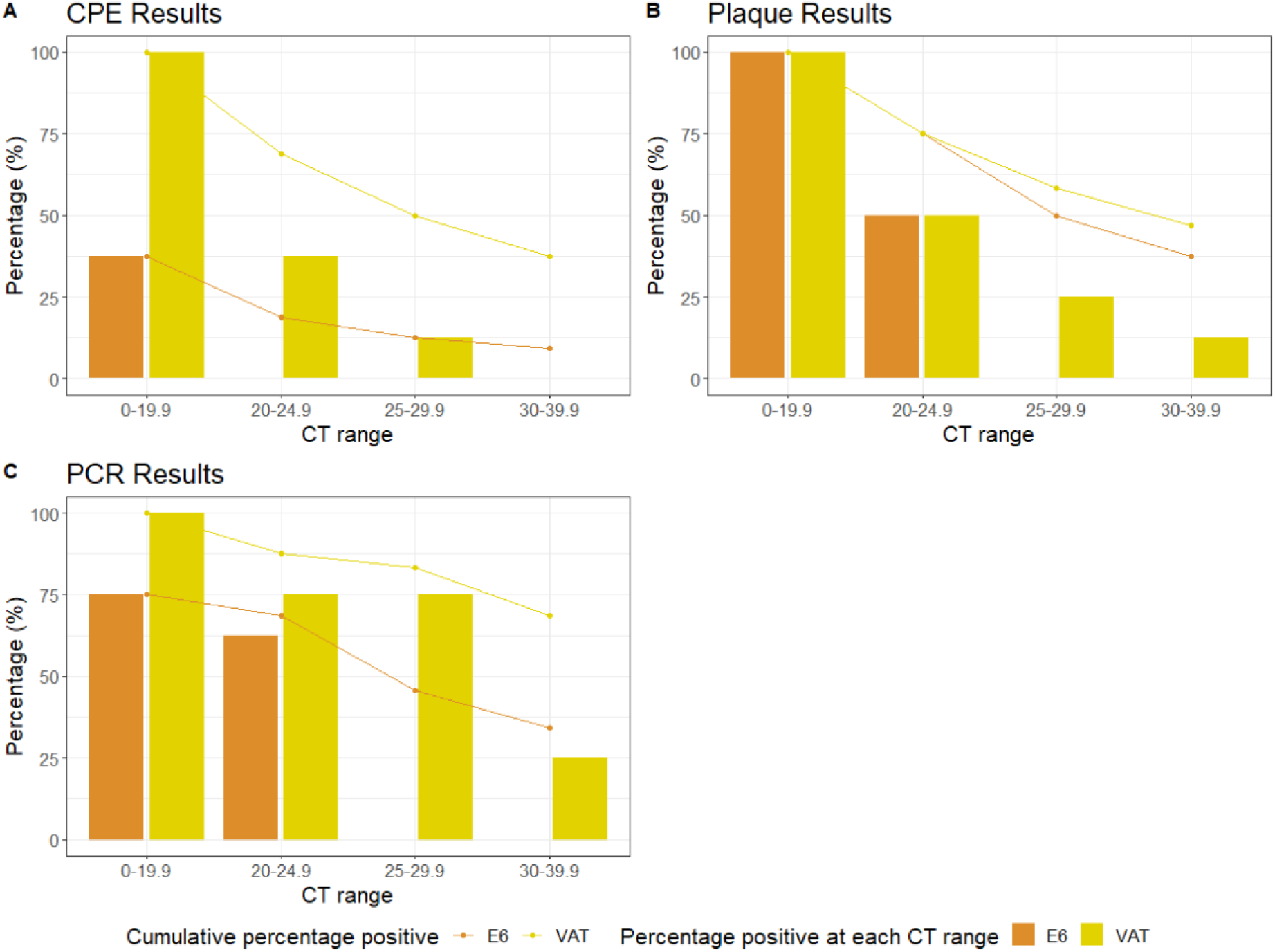
The percentage of positive results according to the different Ct ranges in viral passage 1 using the three detection methods. A: cytopathic effect (CPE), B: Plaque assay and C: RT-qPCR.

Plaque assays from culture supernatants identified viable virus in all samples with a Ct <20 cycles in both cell lines (Fig.4B). This reduced to 50% in both cell lines for the 20-24.9 Ct range. Cumulative positivity rates for each cell line for samples <Ct 30 were 50% for E6 cells and 58.3% for VAT cells. RT-qPCR was the most sensitive detection method for passage 1 (Fig. 4C), particularly when using the VAT cells, which had a cumulative positivity rate of 68.75%, 83.3% 87.5%, and 100%, at Ct ranges of and <40, <30, <25, and <20, respectively.

The VAT cells were the best performing cell line in terms of sensitivity when using Delta variant-containing samples with lower viral loads (i.e., higher Ct values), with two samples with a Ct >30 identified when RT-qPCR was used as a detection method (sample Cts = 31.79, 30.20) (Fig. 5A). The sample with the highest Ct detected by Vero E6 from passage 1 was Ct 25.53. The Vero E6 cells with RT-qPCR combination failed to identify two samples below Ct 20, with Cts of 12.74 and 14.97.

**Figure 5.**
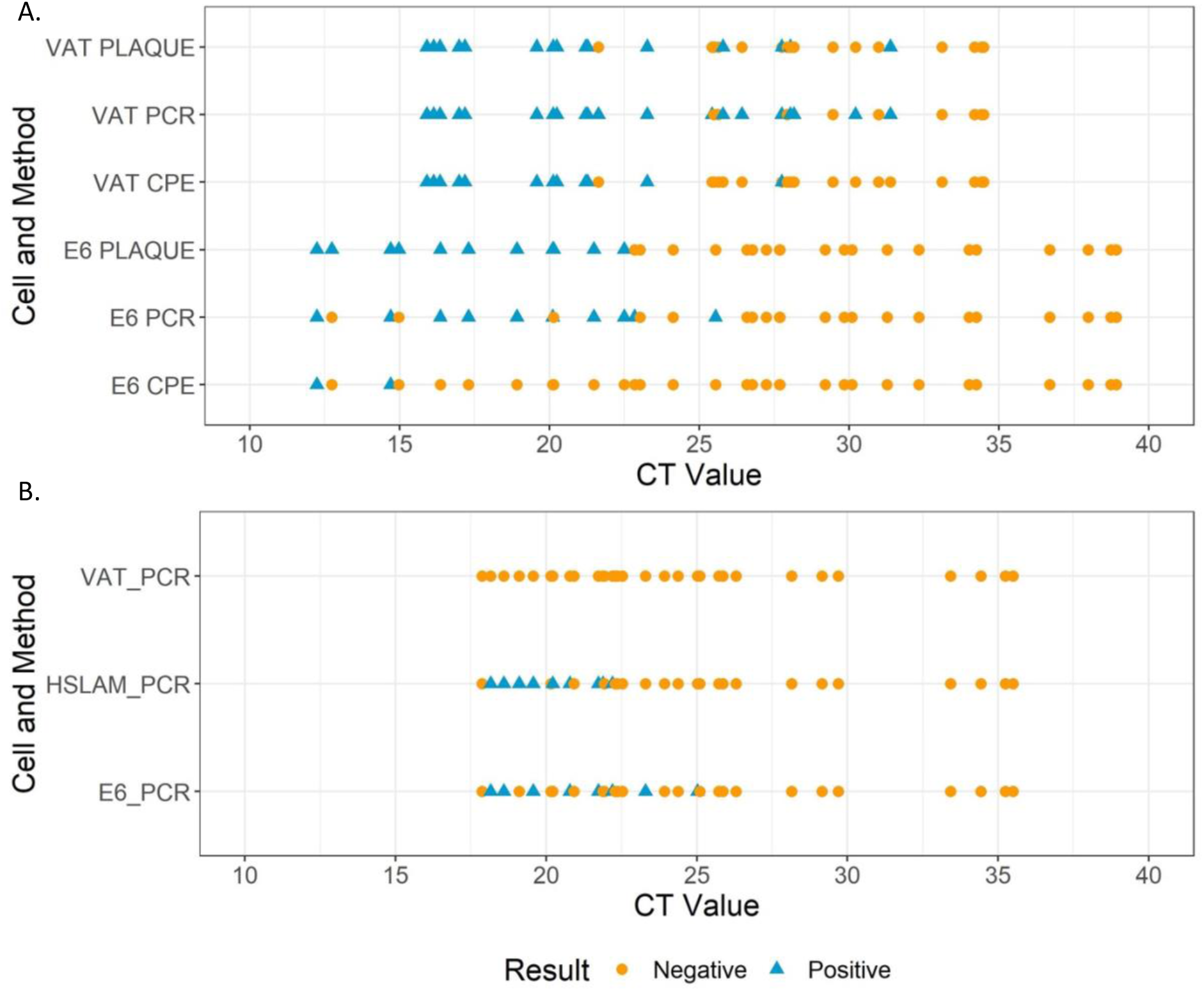
Positive and negative results by original Ct values using the different detection methods with A. Delta and B. Omicron samples, respectively. Each dot or triangle represents an individual sample. Most positive results were identified when samples had Ct<25 regardless of the variant used. CPE: cytopathic effect, Plaque: plaque assay and PCR: RT-qPCR.

Two samples with Ct < 20 (6.25%) assessed with VAT cells failed to produce positive results when microscopy was used as the detection method (Table S3A). The negative control samples were found to be negative by all assays (Table S4), apart from RT-qPCR from VAT cells (1/4 false positive at P3).

Using the final methodology (10µL sample volume, 1 passage of 3 days with VAT cells and RT-qPCR as a detection method) we observed some variation within replicates, particularly with samples with higher Ct values. Of the samples with at least one positive replicate, 67.9% were positive in all replicates, 10.7% in two and 21.4% in one (Table S5).

### Validation of the final culture protocol using Omicron clinical samples

The optimal protocol from the assay development experiments was determined to be a single three-day incubation of samples with a 1:20 dilution, and RT-qPCR as the detection method. This was then assessed using thirty-one positives and 16 negative samples collected during the Omicron outbreak. Three different cell lines: Vero E6, VAT and hSLAM cells were tested using the same sample set in all cases to assess the optimal cell line for Omicron detection.

### RT-qPCR analysis of viral P1 samples

Detection of viable virus was demonstrated to be variable across the three cell lines when assessing passaged clinical samples. Using the optimized protocol developed with clinical samples from the Delta wave, the results showed that both hSLAM and Vero E6 cells resulted in a higher proportion of culture positive samples than the VAT cells (Fig. 6), which were unable to isolate replicative virus from any of the samples tested. From the 31 Omicron positives samples used in the study, 7 failed to give a positive result by RT-qPCR in the baseline pre-culture sample after dilution and were therefore treated as negatives. Of the remaining samples, 8/24 (33.3%), 9/24 (37.5%) and 0/24 (0%) were positive when cultured with Vero E6, hSLAM and VAT cells, respectively.

**Figure 6.**
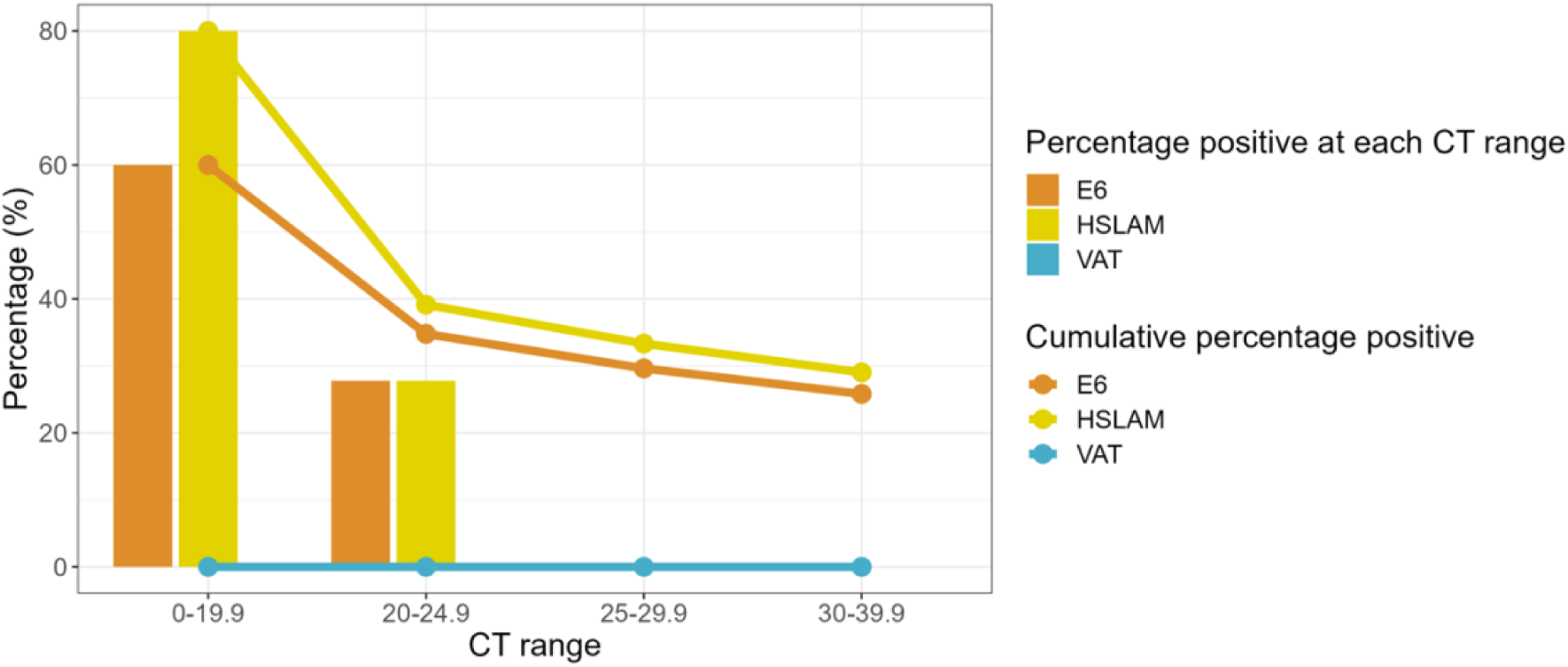
Percentage of Omicron positive results detected by RT-qPCR according to Ct range. Positive results are presented based on the four different Ct categories for each cell line. Solid coloured lines show the cumulative percentage values for each category.

Overall, culture positivity with the hSLAM and Vero E6 cells was 80% and 60% respectively, from samples with a Ct <20 (Fig. 6). In samples with Ct <25, positivity was 35-40% (Fig 5B); no sample with Ct >25 was culture positive in any cell line. (Table S3B). The RT-qPCR negative clinical samples were all found to be negative with the optimized culture protocol (13/13). The positive control (replicating authenticated virus) showed a mean Ct of 17.89, 11.67, and 15.60 for Vero E6, hSLAM and VAT cells when cultured, respectively (Table S6). The negative control and no virus controls were negative regardless of the cell line.

Again, variation was seen within replicates, particularly with samples with higher Ct values. The best performing cell line, hSLAM, had 69.2% of samples positive in all replicates (Table S5).

## DISCUSSION

The use of cell culture as a virological end point in trials of SARS-CoV-2 therapeutics has been limited compared to molecular approaches, ostensibly due to technical difficulties, lack of standardisation, availability of biosafety level 3 laboratories (BSL3) and reduced sensitivity. However, utilising a culture system to monitor replicating virus could mitigate the potential of molecular methods detecting viral RNA from inactive or lysed viral particles.

Here, we have demonstrated that a combination of VAT and hSLAM cells for viral isolation with RT-qPCR of the culture supernatant at passage one for detection is the most sensitive approach for determining the presence of infectious virus of Delta and Omicron VOC, respectively. Culture positivity was higher overall for samples containing the Delta VOC than Omicron, particularly for samples with an RT-qPCR Ct >25.

A previous clinical trial of the antiviral drug Molnupiravir utilised a cell culture/supernatant RT-qPCR approach, with Vero E6 cells, and was able to identify replication-competent virus in 43.5% of infected participants at enrolment, a significant difference between control and treatment groups at day 3, plus a dose response relationship between the drug and viral isolation^5^. In the case of clinical samples for Delta VOC, we found the VAT cells to be more sensitive than Vero E6 cells for isolating SARS-CoV-2, particularly from samples with lower viral loads. It is generally regarded that the ACE2 receptor expressed by the VAT cells is critical for the entry of SARS-CoV-2 to human epithelial cells, with the serine protease TMPRSS2 priming the S protein for binding^31^. In 2020 the expression of TMPRSS2 by Vero E6 cells has been shown to enhance the isolation of wild-type SARS-CoV-2 from clinical samples^31^, which was also apparent in our results with the Delta variant. However, the lack of Omicron viral replication in any clinical sample with the VAT cells was surprising as there was successful viral infection in VAT cells during the growth curve experiments probably due to cell adaptation (albeit with virus passaged through VAT cells previously), and other studies show Omicron can replicate in VAT cells^32,33^. This experiment took place in parallel with the other cell lines, and was further repeated to confirm these findings, with the same results obtained. In contrast, we found that Vero E6 and hSLAM cells were much more effective at isolating virus from Omicron samples. Omicron has low efficiency for TMPRSS-2 mediated cell entry and preferentially infects via cathepsin-mediated endocytosis^29,32^. In addition, it was recently demonstrated that TMPRSS2 activity on both ACE2 and SARS-CoV-2 spike activation is what leads to the significant change in entry requirements from Delta to Omicron lineages ^34^.

This study focussed on viral isolation in Vero cells and their derivatives and did not assess other cell lines reported to be useful culture systems for SARS-CoV-2. Whilst human epithelium-derived cell lines such as Calu-3 and Caco-2 have been used to isolate and propagate SARS-CoV-2, they produce lower viral titres, do not undergo visible CPE or reproducibly allow viral plaques^35^, and are less efficient at viral isolation than Vero E6 cells^36,37^.

The use of RT-qPCR to monitor a change in Ct value was found to be more sensitive than microscopy or plaque assay, enabling the detection of replicative virus in the absence of overt CPE. The use of a Ct difference of >1 Ct for culture positivity was selected based on the reproducibility of the assay, and the prediction that RNA from non-replicative virus would decay during incubation, however an increased Ct difference could be selected to increase specificity whilst potentially reducing sensitivity. Due to failed controls on plates (e.g., the virus free control having a disrupted monolayer making result interpretation impossible), the plaque assay approach produced a substantial proportion of unsuccessful replicates, resulting in the need for repeats. While RT-qPCR is the most sensitive and rapid analytical technique used, the data generated require longitudinal analysis to demonstrate the presence of replicating virus.

The methodologies reported utilised 24-well microtiter plates to provide a reasonable throughput whilst enabling sufficient inoculum volume to maintain sensitivity. These assays could potentially be carried out in 48 or 96 well plates to maximise throughput or scaled up to 6 well plates or flasks to maximise input volume and potentially sensitivity. Whilst we added antibiotics and antimycotics to cell culture media to reduce contamination, filtering the inoculum could lessen the need for inoculum dilution thereby benefitting sensitivity. Other culture-based methods such as TCID_50_ assays can be done at a higher throughput than plaque assays, however these were not evaluated during our method development.

The specificity of the assay was found to be high, with the only false positives found in passage 3 by RT-qPCR for Delta and no false positives for Omicron. This could have been due to contamination of the cell culture or RT-qPCR assays. The study only included four SARS-CoV-2 negative swab samples for Delta analysis and further testing is required to assess the test specificity more confidently. In the case of analysis against Omicron clinical samples, a greater number of negative samples were included (n=16).

Despite lower sensitivity than RT-qPCR at detecting the presence of SARS-CoV-2 in clinical samples, the greater specificity of cell culture, which only detects viable virus, improves the ability to evaluate efficacy of an antiviral by revealing a larger difference between the treatment and control arms. Infection and viral load kinetics differ between SARS-CoV-2 variants, and this work reinforces the need to verify cell line suitability for circulating variants before the selection of cell line for culture-based diagnostics. Even though more effort and caution are required as variations alter, the improved data produced may benefit research projects such as clinical trials.

Here we have optimised a cell culture-based assay for determining the presence of infectious SARS-CoV-2 Delta and Omicron variants using clinical nasopharyngeal swabs, determining the optimal sample dilution, culture time, cell line, passage, and detection method. We have also identified an ongoing need to periodically re-assess optimal cell lines throughout the pandemic as the virus evolves and receptor usage and tropism changes over time. This methodology may have application as a secondary virological end point in clinical trials of therapeutics for SARS-CoV-2 in addition to numerous research processes.

## Author contributions

BA, TF and TE conceptualised the study. RW, DB, KB, DW, HS, ACA contributed to sample collection during the FALCON study. DW, KB, ARR, VS, TF, and TE contributed to the experimental design and data analysis. DW, KB, ARR, RW, DB, CGB, CT, NK and HS carried out laboratory experiments. Data analysis was carried out by DW, KB, ARR, HS and TE. Funding was received by BG, ERA, SK, TF and TE. DW, KB, ARR, HS, TF and TE wrote the first draft of the manuscript. All authors reviewed and edited the final manuscript.

## Data Availability

All data produced in the present work are contained in the manuscript

## Acknowledgements

Professor Wendy Barclay (Imperial College London) kindly provided the VAT cells. We also wish to acknowledge the CONDOR steering group who lead the FALCON study from which our samples were taken.

We acknowledge the support of the National Institute for Health Research (NIHR) Clinical Research Network, which supports delivery of the FALCON study. The views expressed in this article are those of the authors and not necessarily those of the NIHR, or the Department of Health and Social Care.

Condor steering group: Dr A. Joy Allen, Dr Julian Braybrook, Professor Peter Buckle, Professor Paul Dark, Dr Kerrie Davis, Professor Adam Gordon, Ms Anna Halstead, Dr Charlotte Harden, Dr Colette Inkson, Ms Naoko Jones, Dr William Jones, Professor Dan Lasserson, Dr Joseph Lee, Dr Clare Lendrem, Dr Andrew Lewington, Mx Mary Logan, Dr Massimo Micocci, Dr Brian Nicholson, Professor Rafael Perera-Salazar, Mr Graham Prestwich, Dr D. Ashley Price, Dr Charles Reynard, Dr Beverley Riley, Professor AJ Simpson, Dr Valerie Tate, Dr Philip Turner, Professor Mark Wilcox, Dr Melody Zhifang.

## Supplementary List

1. **Methods**

a. Sample dilution
b. SARS-CoV-2 growth curves
2. Results

a. Viral growth kinetics for Delta
b. Viral growth kinetics for Omicron

**Figure S1.** Growth curve of the A. Delta and B. Omicron isolate.

**Figure S2.** RT-qPCR Ct values of Delta clinical samples tested by each cell line.

**Table S1.** Optimisation of the sample input volume.

**Table S2.** RT-qPCR and plaque assay results per cell line for each day with Delta and Omicron viral stocks.

Table S3. Total results by cell line for each passage with A. Delta and B. Omicron samples.

**Table S4**. Results using Delta negative control samples.

**Table S5.** Variation across replicates during the RT-qPCR method in samples with at least one positive replicate.

**Table S6.** Results of the Omicron controls used along with the clinical samples.

## REFERENCES

1 Sakamaki, K., Uemura, Y. & Shimizu, Y. Definitions and elements of endpoints in phase III randomized trials for the treatment of COVID-19: a cross-sectional analysis of trials registered in ClinicalTrials.gov. Trials 22, 788 (2021). 10.1186/s13063-021-05763-y

2 Desai, A. & Gyawali, B. Endpoints used in phase III randomized controlled trials of treatment options for COVID-19. 23, 100403 (2020).

3 Mehta, H. B., Ehrhardt, S., Moore, T. J., Segal, J. B. & Alexander, G. C. Characteristics of registered clinical trials assessing treatments for COVID-19: a cross-sectional analysis. BMJ Open 10, e039978 (2020).

4 Wang, Y. et al. Remdesivir in adults with severe COVID-19: a randomised, double-blind, placebo-controlled, multicentre trial. 395, 1569–1578 (2020).

5 Fischer William, A., et al. A phase 2a clinical trial of molnupiravir in patients with COVID-19 shows accelerated SARS-CoV-2 RNA clearance and elimination of infectious virus. Science Translational Medicine 14, eabl7430

6 Hammond, J. et al. Oral Nirmatrelvir for High-Risk, Nonhospitalized Adults with Covid-19. N Engl J Med 386, 1397–1408 (2022).

7 Kevadiya, B. D. et al. Diagnostics for SARS-CoV-2 infections. 20, 593–605 (2021).

8 Singanayagam, A. et al. Duration of infectiousness and correlation with RT-PCR cycle threshold values in cases of COVID-19, England, January to May 2020. Eurosurveillance 25, 2001483 (2020). 10.2807/1560-7917.ES.2020.25.32.2001483

9 van Kampen, J. J. A. et al. Duration and key determinants of infectious virus shedding in hospitalized patients with coronavirus disease-2019 (COVID-19). 12, 267 (2021).

10 Cevik, M. et al. SARS-CoV-2, SARS-CoV, and MERS-CoV viral load dynamics, duration of viral shedding, and infectiousness: a systematic review and meta-analysis. The lancet microbe 2, e13–e22 (2021).

11 Wurtz, N., Penant, G., Jardot, P., Duclos, N. & La Scola, B. Culture of SARS-CoV-2 in a panel of laboratory cell lines, permissivity, and differences in growth profile. European Journal of Clinical Microbiology & Infectious Diseases 40, 477–484 (2021). 10.1007/s10096-020-04106-0

12 Wei, J. et al. Genome-wide CRISPR screens reveal host factors critical for SARS-CoV-2 infection. Cell 184, 76–91. e13 (2021).

13 Murgolo, N. et al. SARS-CoV-2 tropism, entry, replication, and propagation: Considerations for drug discovery and development. PLOS Pathogens 17, e1009225 (2021). 10.1371/journal.ppat.1009225

14 Leonard, V. H. J., Hodge, G., Reyes-Del Valle, J., McChesney, M. B. & Cattaneo, R. Measles virus selectively blind to signaling lymphocytic activation molecule (SLAM; CD150) is attenuated and induces strong adaptive immune responses in rhesus monkeys. Journal of virology 84, 3413–3420 (2010).

15 Rihn, S. J. et al. A plasmid DNA-launched SARS-CoV-2 reverse genetics system and coronavirus toolkit for COVID-19 research. PLOS Biology 19, e3001091 (2021).

16 Prince, T. et al. Analysis of SARS-CoV-2 in Nasopharyngeal Samples from Patients with COVID-19 Illustrates Population Variation and Diverse Phenotypes, Placing the Growth Properties of Variants of Concern in Context with Other Lineages. mSphere, e0091321 (2022). 10.1128/msphere.00913-21

17 Matsuyama, S. et al. Enhanced isolation of SARS-CoV-2 by TMPRSS2-expressing cells. Proceedings of the National Academy of Sciences 117, 7001–7003 (2020). 10.1073/pnas.2002589117

18 Folgueira, M. D., Luczkowiak, J., Lasala, F., Pérez-Rivilla, A. & Delgado, R. Prolonged SARS-CoV-2 cell culture replication in respiratory samples from patients with severe COVID-19. Clinical microbiology and infection : the official publication of the European Society of Clinical Microbiology and Infectious Diseases 27, 886–891 (2021).

19 Berengua, C. et al. Viral culture and immunofluorescence for the detection of SARS-CoV-2 infectivity in RT-PCR positive respiratory samples. Journal of clinical virology : the official publication of the Pan American Society for Clinical Virology 152, 105167–105167 (2022).

20 Mendoza, E. J., Manguiat, K., Wood, H. & Drebot, M. Two Detailed Plaque Assay Protocols for the Quantification of Infectious SARS-CoV-2. Curr Protoc Microbiol 57, ecpmc105–ecpmc105 (2020). 10.1002/cpmc.105

21 Craig, N. et al. Direct Lysis RT-qPCR of SARS-CoV-2 in Cell Culture Supernatant Allows for Fast and Accurate Quantification. Viruses 14, 508 (2022).

22 Biosystems, A. TaqPath™ COVID-19 Combo Kit INSTRUCTIONS FOR USE. (2020).

23 Tchesnokova, V. et al. Acquisition of the L452R mutation in the ACE2-binding interface of Spike protein triggers recent massive expansion of SARS-Cov-2 variants. Journal of clinical microbiology 59, e00921–00921 (2021).

24 Hodcroft, E. e. a. CoVariants-Overview of Variants/Mutations, <https://covariants.org/per-variant> (

25 Li, A., Maier, A., Carter, M. & Guan, T. H. Omicron and S-gene target failure cases in the highest COVID-19 case rate region in Canada—December 2021. Journal of medical virology 94, 1784 (2022).

26 Metzger, C. M. et al. PCR performance in the SARS-CoV-2 Omicron variant of concern? Swiss medical weekly 151, w30120–w30120 (2021).

27 Patterson, E. I. et al. Methods of inactivation of SARS-CoV-2 for downstream biological assays. The Journal of infectious diseases 222, 1462–1467 (2020).

28 Pawar, S. D. et al. Replication of SARS-CoV-2 in cell lines used in public health surveillance programmes with special emphasis on biosafety. Indian Journal of Medical Research 155, 129–135 (2022).

29 Dighe, H. et al. Differential cell line susceptibility to the SARS-CoV-2 omicron BA. 1.1 variant of concern. Vaccines 10, 1962 (2022).

30 R Core Team, R. R: A language and environment for statistical computing. (2013).

31 Hoffmann, M. et al. SARS-CoV-2 Cell Entry Depends on ACE2 and TMPRSS2 and Is Blocked by a Clinically Proven Protease Inhibitor. Cell 181, 271–280.e278 (2020). 10.1016/j.cell.2020.02.052

32 Meng, B. et al. Altered TMPRSS2 usage by SARS-CoV-2 Omicron impacts infectivity and fusogenicity. Nature 603, 706–714 (2022).

33 Saito, A. et al. Virological characteristics of the SARS-CoV-2 Omicron BA. 2.75 variant. Cell host & microbe 30, 1540–1555. e1515 (2022).

34 Aggarwal, A. et al. TMPRSS2 activation of Omicron lineage Spike glycoproteins is regulated by TMPRSS2 cleavage of ACE2. bioRxiv, 2023.2009. 2022.558930 (2023).

35 Mautner, L. et al. Replication kinetics and infectivity of SARS-CoV-2 variants of concern in common cell culture models. 19, 76 (2022).

36 Essaidi-Laziosi, M. et al. Estimating clinical SARS-CoV-2 infectiousness in Vero E6 and primary airway epithelial cells. The Lancet Microbe 2, e571 (2021).

37 Funnell, S. G. et al. A cautionary perspective regarding the isolation and serial propagation of SARS-CoV-2 in Vero cells. NPJ vaccines 6, 83 (2021).

