## Supplementary materials for "Optimisation of SARS-CoV-2 culture from clinical samples for clinical trial applications"

### Supplementary material

#### Methods

##### a. Sample dilution

To determine the optimal dilution factor, clinical samples previously tested negative for SARS-CoV-2 by RT-qPCR (TaqPath COVID-19 RT-qPCR on QuantStudio 5 (ThermoFisher, USA) were spiked with $3x10^4$ pfu/ml of SARS-CoV-2 virus Delta VOC (to give easily visible plaques without destroying the monolayer). Media was removed from confluent 24-well plates. Wells were inoculated with either 10µL, 20µL or 50µL of the spiked negative clinical sample added to 190µL, 180µL or 150µL of D2 media. Plates were incubated at 37°C + 5% $CO_2$ for 2 hours, before 200 µL/well of D10 media was added. Plates were then returned to the incubator for 72 hours, after which they were checked for contamination both visually and using a microscope. The process was repeated twice, with the same volume transferred to fresh confluent 24-well plates with appropriate volumes of D2 media, as above. The remaining volume from each well was transferred to cryotubes and stored at -80$^o$C. Once viral culture had been collected from all three passages, samples were thawed, and plaque assays were performed as described below. The criteria to determine positive samples was the presence of plaques otherwise was determined as negative. The dilution that consistently produced plaques without causing fungal or bacterial contamination was carried forwards into the further experiments.

##### b. SARS-CoV-2 growth curves

To determine the optimal number of days growth of virus per passage, media was removed from a pre-prepared 24-well plate seeded with each cell line for Delta (Genbank accession number: (SARS-CoV-2/human/GBR/Liv_273/2021) and Omicron (BA.1) (SARS-CoV-2/human/GBR/Liv_1326/2021), and 190µL of D2 media was added with 10µL/well of a $3x10^4$ pfu/mL of SARS-CoV-2. Plates were incubated at 37°C + 5% $CO_2$ for 2 hours, before 200µL of D10 media was added, and plates returned to the incubator. Aliquots of 140µL from two wells were taken at 24, 48, 72, 96 and 120 hours, and stored at -80°C. Samples were thawed and tested using both plaque assays and RT-qPCR, performed as described in Section 3 and 4 from Methods.

#### Results

##### Viral growth kinetics for Delta

Both replicative virus and viral RNA were detectable in each cell line at day one, with viral loads then increasing from day two to three. Viral RNA levels then plateaued between day three and five, whilst viable virus declined (Fig. S1A, Table S2). The highest viral load at days one and two were found in the VAT cells; however, by day three both cell lines had equivalent viral loads (mean Ct difference 0.59). Therefore, a three-day passage was chosen to obtain high viral loads whilst minimising culture time.

**Viral growth kinetics for Omicron**

Viral growth was also measured using the Omicron viral stock for over a 5-day period in three different cell lines. The growth kinetics of Omicron infection revealed comparable curves in hSLAM and VAT cells lines, with lower viral loads in Vero E6 (Fig. 2B). The viral load increased with time regardless of the cell line, peaking at day 5 post-infection with mean Ct values of 17.84, 12.02 and 9.92 for Vero E6, hSLAM and VAT cells, respectively (Table S2). The use of hSLAM resulted in higher viral RNA levels, which plateaued between day 3 and 4 and increased at day 5 post infection. Lower titres of PFU and RNA were detected from cultures in Vero E6.

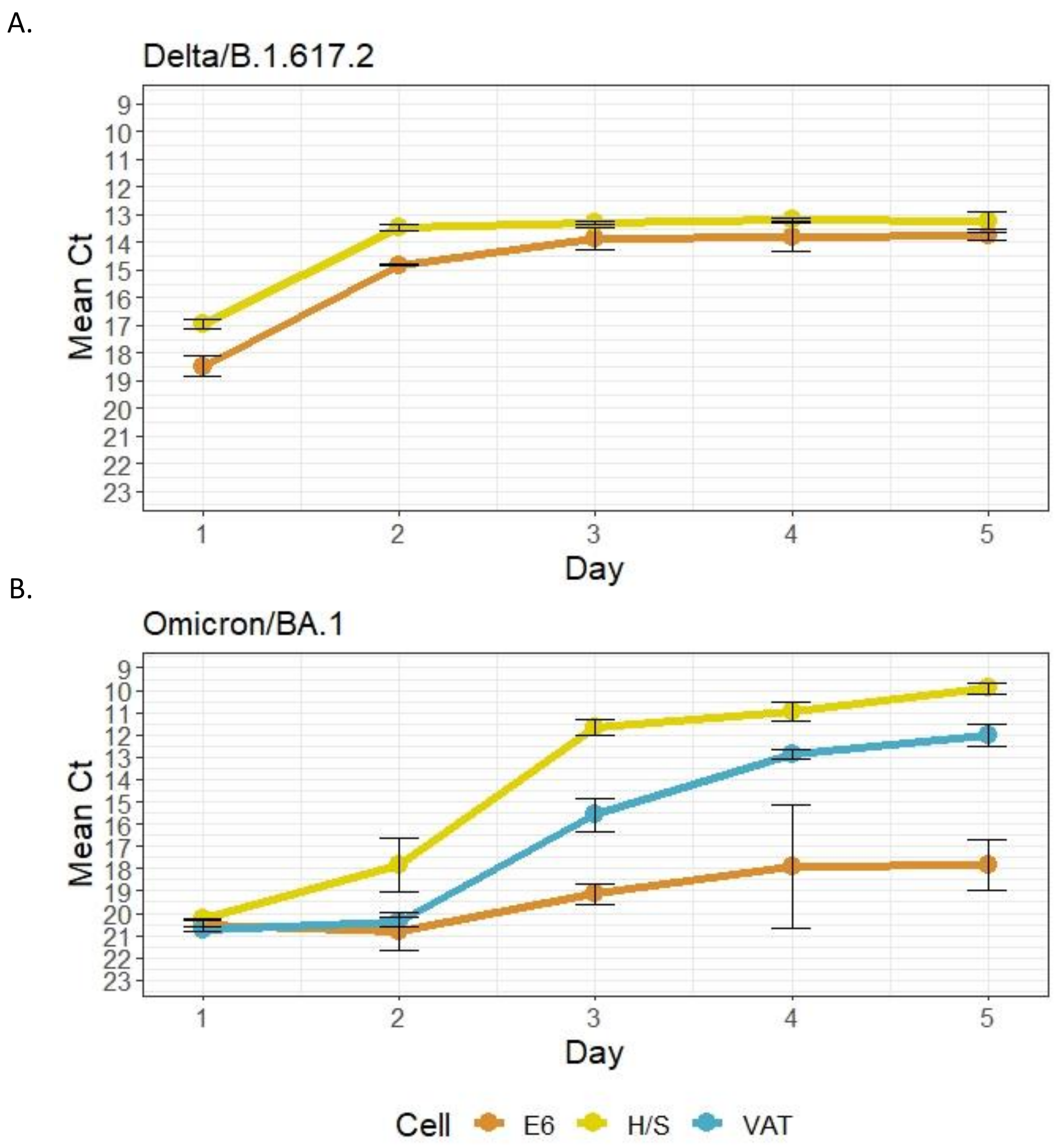


**Figure S1.** Growth curve of the A. Delta and B. Omicron isolate. Result of RT-qPCR representing the mean Ct values during a 5-day period incubation in both Vero E6, VAT and hSLAM cells. In the case of Omicron, while Ct values in Vero E6 cells increased at day 5, VAT and hSLAM cells continued to reduce the viral load throughout time. E6: VERO-E6 cells, H/S: hSLAM cells.

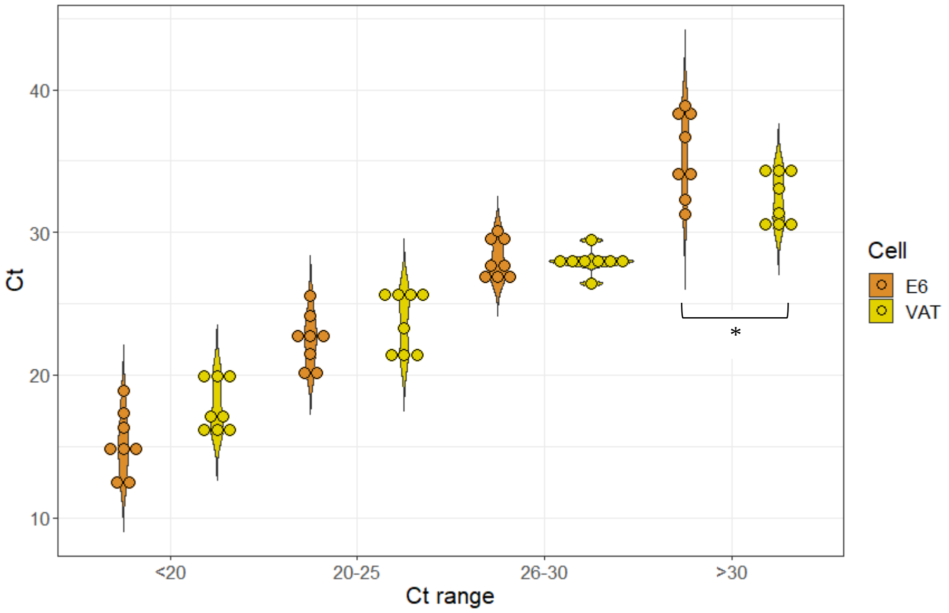


**Figure S2.** RT-qPCR Ct values of Delta clinical samples tested by each cell line. The mean Ct is represented for each cell line at a specific Ct range. Clinical samples cultured in Vero E6 showed a significantly higher mean Ct compared to samples cultured in VAT cells in the Ct>30 range (marked by an asterisk, p-value= 0.025). The other Ct ranges showed no significant differences between cell lines.

**Table S1.** Optimisation of the sample input volume. Wells with plaques (P) and wells contaminated (C) were counted for each passage (P1, P2 and P3) in both Vero E6 and VAT infected cell lines.

| **Volume (µL)** | **Vero E6** | | | **VAT** | | |
|---|---|---|---|---|---|---|
| | **P1** | **P2** | **P3** | **P1** | **P2** | **P3** |
| 10 µL | P: 3/3<br>C: 0/3 | P: 3/3<br>C: 0/3 | P: 3/3<br>C: 0/3 | P: 3/3<br>C: 0/3 | P: 3/3<br>C: 0/3 | P: 3/3<br>C: 0/3 |
| 20 µL | P: 3/3<br>C: 0/3 | P: 3/3<br>C: 1/3 | P: 3/3<br>C: 1/3 | P: 3/3<br>C: 0/3 | P: 3/3<br>C: 0/3 | P: 3/3<br>C: 0/3 |

| | | | | | | |
|---|---|---|---|---|---|---|
| 50 µL | P: 3/3<br>C: 2/3 | P: 3/3<br>C: 2/3 | P: 3/3<br>C: 3/3 | P: 3/3<br>C: 0/3 | P: 3/3<br>C: 0/3 | P: 3/3<br>C: 0/3 |

**Table S2**. RT-qPCR and plaque assay results per cell line for each day with Delta and Omicron viral stocks. The results from the RT-qPCR are shown as mean Ct values and the number of plaques is represented in pfu/mL. P1, P2 and P3 refers to passage 1, 2, and 3, respectively.

| | **Delta** | | | | **Omicron** | | |
|---|---|---|---|---|---|---|---|
| | **Vero E6** | | **VAT** | | **Vero E6** | **h/SLAM** | **VAT** |
| Day | RT-qPCR | Plaque | RT-qPCR | Plaque | RT-qPCR | RT-qPCR | RT-qPCR |
| 1 | 18.47 | $1.5x10^5$ | 16.97 | $4x10^6$ | 20.36 | 20.27 | 20.72 |
| 2 | 14.82 | $4x10^5$ | 13.48 | $1.6x10^7$ | 19.67 | 17.84 | 20.39 |
| 3 | 13.87 | $1x10^5$ | 13.28 | $1.5x10^6$ | 17.88 | 11.67 | 15.60 |
| 4 | 13.79 | $6x10^1$ | 13.21 | $2.5x10^3$ | 17.99 | 10.95 | 12.89 |
| 5 | 13.78 | $3x10^2$ | 13.22 | $1x10^2$ | 19.56 | 9.92 | 12.02 |

**Table S3.** Total results by cell line for each passage with A. Delta and B. Omicron samples. h/SLAM cells showed the highest percentage of positive samples followed by VERO E6 cells with 29.03 and 25.80%, respectively. Supernatants recovered from VAT cells showed 0% positivity contrary to the results obtained with Delta samples.

A.

| Cell type | Result | Passage 1 | | | Passage 2 | | | Passage 3 | | |
|---|---|---|---|---|---|---|---|---|---|---|
| | | CPE | Plaque | PCR | CPE | Plaque | PCR | CPE | Plaque | PCR |
| Vero E6 | Total positive | 3 | 12 | 11 | 11 | 13 | 14 | 16 | 18 | 14 |
| | Total negative | 29 | 20 | 21 | 21 | 19 | 18 | 16 | 14 | 18 |
| | Total failure | 0 | 0 | 0 | 0 | 0 | 0 | 0 | 0 | 0 |
| | Percentage positive | 9.38 | 37.5 | 34.38 | 34.38 | 40.63 | 43.75 | 50 | 56.25 | 43.75 |
| | Percentage negative | 90.63 | 62.5 | 65.63 | 65.63 | 59.38 | 56.25 | 50 | 43.75 | 56.25 |
| | Percentage culture negative | 0 | 0 | 0 | 0 | 0 | 0 | 0 | 0 | 0 |
| VAT | Total positive | 12 | 15 | 22 | 10 | 11 | 12 | 9 | 13 | 13 |
| | Total negative | 20 | 17 | 10 | 20 | 21 | 20 | 21 | 19 | 19 |
| | Total failure | 0 | 0 | 0 | 2 | 0 | 0 | 2 | 0 | 0 |
| | Percentage positive | 37.5 | 46.88 | 68.75 | 31.25 | 34.38 | 37.5 | 28.13 | 40.63 | 40.63 |
| | Percentage negative | 62.5 | 53.13 | 31.25 | 62.5 | 65.63 | 62.5 | 65.63 | 59.38 | 59.38 |
| | Percentage culture negative | 0 | 0 | 0 | 6.25 | 0 | 0 | 6.25 | 0 | 0 |

B.

| Result | Vero E6 | h/SLAM | VAT |
|---|---|---|---|
| Total positive | 8 | 9 | 0 |
| Total negative | 16 | 15 | 24 |
| Total failure | 7 | 7 | 7 |
| Percentage positive | 25.80 | 29.03 | 0 |
| Percentage negative | 51.61 | 48.39 | 77.42 |
| Percentage culture negative | 22.58 | 22.58 | 22.58 |

| Cell type | Result | P1 | | | P2 | | | P3 | | |
|---|---|---|---|---|---|---|---|---|---|---|
| | | CPE | Plaque | PCR | CPE | Plaque | PCR | CPE | Plaque | PCR |
| Vero E6 | >40 - Undetermined | 0 | 0 | 0 | 0 | 0 | 0 | 0 | 0 | 0 |
| | >40 - Undetermined | 0 | 0 | 0 | 0 | 0 | 0 | 0 | 0 | 0 |
| | >40 - Undetermined | 0 | 0 | 0 | 0 | 0 | 0 | 0 | 0 | 0 |
| | >40 - Undetermined | 0 | 0 | 0 | 0 | 0 | 0 | 0 | 0 | 0 |
| | Total positive | 0 | 0 | 0 | 0 | 0 | 0 | 0 | 0 | 0 |
| | Total negative | 4 | 4 | 4 | 4 | 4 | 4 | 4 | 4 | 4 |
| | Failure | 0 | 0 | 0 | 0 | 0 | 0 | 0 | 0 | 0 |
| | Percentage positive | 0 | 0 | 0 | 0 | 0 | 0 | 0 | 0 | 0 |
| | Percentage negative | 100 | 100 | 100 | 100 | 100 | 100 | 100 | 100 | 100 |
| | Percentage Failure | 0 | 0 | 0 | 0 | 0 | 0 | 0 | 0 | 0 |
| VAT | >40 - Undetermined | 0 | 0 | 0 | 0 | 0 | 0 | 0 | 0 | 0 |
| | >40 - Undetermined | 0 | 0 | 0 | 0 | 0 | 0 | 0 | 0 | 0 |
| | >40 - Undetermined | 0 | 0 | 0 | 0 | 0 | 0 | 0 | 0 | 0 |
| | >40 - Undetermined | 0 | 0 | 0 | 0 | 0 | 0 | 0 | 0 | 1 |
| | Total positive | 0 | 0 | 0 | 0 | 0 | 0 | 0 | 0 | 1 |
| | Total negative | 4 | 4 | 4 | 4 | 4 | 4 | 4 | 4 | 3 |
| | Failure | 0 | 0 | 0 | 0 | 0 | 0 | 0 | 0 | 0 |
| | Percentage positive | 0 | 0 | 0 | 0 | 0 | 0 | 0 | 0 | 25 |
| | Percentage negative | 100 | 100 | 100 | 100 | 100 | 100 | 100 | 100 | 75 |
| | Percentage Failure | 0 | 0 | 0 | 0 | 0 | 0 | 0 | 0 | 0 |

**Table S4.** Results using Delta negative control samples.

| Result | Delta | | Omicron | | |
|---|---|---|---|---|---|
| | Vero E6 | VAT | Vero E6 | h/SLAM | VAT |
| 3/3 Positive | 5/13 (38.5%) | 19/28 (67.9%) | 8/13 (62.5%) | 9/13 (69.23%) | 0/2 (0%) |
| 2/3 Positive | 6/13 (46.2%) | 3/28 (10.7%) | 0/13 (0%) | 0/13 (0%) | 0/2 (0%) |
| 1/3 Positive | 2/13 (15.4%) | 6/28 (21.4%) | 5/13 (38.46%) | 4/13 (30.77%) | 2/2 (100%) |

**Table S5.** Variation across replicates during the RT-qPCR method in samples with at least one positive replicate. hSLAM and Vero E6 cell lines showed the best results with high percentage of positive samples while supernatants collected from VAT cells demonstrated the lowest positivity percentage of all cell lines.

| Control | VERO E6 | H/SLAM | VAT |
|---|---|---|---|
| Positive control (Live virus) | 17.89 | 11.67 | 15.6 |
| Negative control (inactivated virus) | 38.02 | 33.95 | 37.24 |
| Blank (UTM only) | ND | ND | ND |

**Table S6.** Results of the Omicron controls used along with the clinical samples. Results showed high viral load on the positive controls regardless the cell line, while the negative controls showed very low viral loads. The was no RNA detected on the blank control samples. ND: Non detectable.
